# Molecular analysis for ovarian cancer detection in patient-friendly samples

**DOI:** 10.1101/2023.09.28.23296279

**Authors:** Birgit M.M. Wever, Mirte Schaafsma, Maaike C.G. Bleeker, Yara van den Burgt, Rianne van den Helder, Christianne A.R. Lok, Frederike Dijk, Ymke van der Pol, Florent Mouliere, Norbert Moldovan, Nienke E. van Trommel, Renske D.M. Steenbergen

## Abstract

**Background:** High ovarian cancer mortality rates motivate the development of effective and patient-friendly diagnostics. Here, we explored the potential of molecular testing in patient-friendly samples for ovarian cancer detection.

**Patients and methods:** Home-collected urine, cervicovaginal self-samples, and clinician-taken cervical scrapes were prospectively collected from 54 patients diagnosed with a highly suspicious ovarian mass (benign n=25, malignant n=29). All samples were tested for nine methylation markers, using quantitative methylation-specific PCRs that were verified on ovarian tissue samples, and compared to unpaired patient-friendly samples of 110 healthy controls. Copy number analysis was performed on a subset of urine samples of ovarian cancer patients by shallow whole-genome sequencing.

**Results:** Three methylation markers were significantly elevated in full void urine of ovarian cancer patients as compared to healthy controls (*C2CD4D*, *p*=0.008; *CDO1*, *p*=0.022; *MAL*, *p*=0.008), of which two were also discriminatory in cervical scrapes (*C2CD4D*, *p*=0.001; *CDO1*, *p*=0.004). When comparing benign and malignant ovarian masses, *GHSR* showed significantly elevated methylation levels in the urine sediment of ovarian cancer patients (*p*=0.024). Other methylation markers demonstrated comparably high methylation levels in benign and malignant ovarian masses. Cervicovaginal self-samples showed no elevated methylation levels in patients with ovarian masses as compared to healthy controls. Copy number changes were identified in 4 out of 23 urine samples of ovarian cancer patients.

**Conclusion:** Our study revealed increased methylation levels of ovarian cancer-associated genes and copy number aberrations in the urine of ovarian cancer patients. Our findings support continued research into urine biomarkers for ovarian cancer detection and highlight the importance of including benign ovarian masses in future studies to develop a clinically useful test.

**HIGHLIGHTS:**

- Ovarian cancer is often diagnosed at an advanced stage with a poor prognosis
- We studied the potential of molecular testing in different types of patient-friendly material for ovarian cancer detection
- Elevated methylation of ovarian cancer-associated genes can be measured in cervical scrapes and urine
- Copy number aberrations are detectable in urine of ovarian cancer patients
- DNA-based testing in cervical scrapes and urine could aid ovarian cancer diagnosis upon further development

**GRAPHICAL ABSTRACT:** 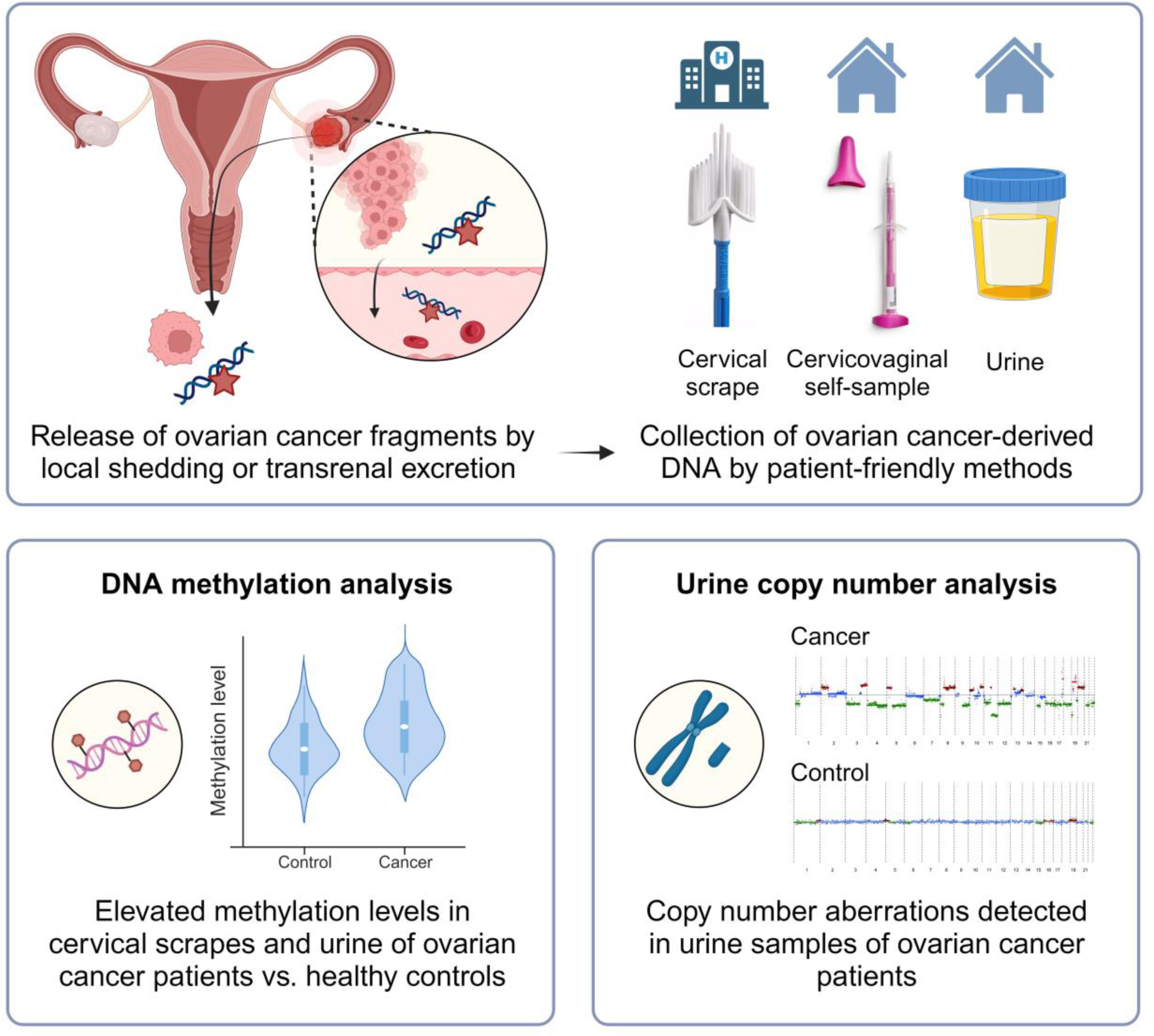

Created with BioRender.com.

## INTRODUCTION

Ovarian cancer is the most lethal gynecological cancer worldwide, accounting for 207.252 deaths in 2020.^1^ Due to non-specific or absence of symptoms at an early-stage, patients typically present at a late-stage when prognosis is poor.^2^ Five-year overall survival rates sharply decrease with higher stage at diagnosis, with 92% survival in early-stage disease compared to only 29% in late-stage disease.^3^ High mortality rates prioritize the development of novel diagnostic approaches for ovarian cancer. Although more ovarian cancer patients were diagnosed at an earlier stage with screening strategies using conventional imaging and/or serum biomarkers (*e.g.,* CA-125), this did not translate into reduced overall cancer-specific mortality in general and in high-risk populations.^4, 5^ In fact, the majority of ovarian cancers were not detected during or after the trial. A more accurate and easily accessible test could potentially overcome this problem.

Testing for ovarian cancer using biomarkers related to carcinogenesis could offer such an accurate test. DNA methylation-mediated silencing of tumor suppressor genes occurs early in cancer development and is therefore promising to detect cancer at an early-stage.^6^ Methylation analysis in urine, cervicovaginal self-samples, and clinician-taken cervical scrapes has already been proven to allow reliable detection of cervical^7, 8^ and endometrial cancer.^9, 10^ In urine, even signals of non-urogenital cancers, including colorectal^11^ and lung cancer^12, 13^, are detectable by methylation testing. The measurement of somatic mutations, aneuploidy, or DNA methylation in clinician-taken cervical scrapes or blood demonstrated the high potential of molecular-based diagnostic tests for ovarian cancer.^14–17^ However, these molecular changes have not been investigated in home-collected urine and cervicovaginal self-samples of ovarian cancer patients.

In this study, we explored the potential of molecular testing in home-collected urine and cervicovaginal self-samples, and clinician-taken cervical scrapes for ovarian cancer detection. Methylation markers considered suitable for the detection of ovarian cancer included a combination of markers described in studies on cervical and endometrial cancer detection in patient-friendly sample types (*GALR1, GHSR, MAL, PRDM14, SST*, and *ZIC1*^10, 18–20^), and ovarian cancer detection in cervical scrapes and plasma (*C2CD4D*, *CDO1*, *NRN1*^17, 21, 22^). In addition, the analysis of somatic copy number aberrations (SCNA) and fragmentation patterns was performed using shallow whole-genome sequencing on a subset of the samples to verify the presence of ovarian cancer-derived DNA in urine.

## MATERIAL AND METHODS

### Study population

This study prospectively included patients with a highly suspicious ovarian mass according to current triage methods (>40% risk of malignancy using the IOTA adnex model).^23, 24^ Paired samples (*i.e.,* urine, cervicovaginal self-samples, and clinician-taken cervical scrapes) were consecutively collected within the SOLUTION1 study, between July 2018 and September 2022, at the Antoni van Leeuwenhoek hospital, Amsterdam, The Netherlands. Samples were collected from patients who underwent pelvic surgery with post-operatively confirmed ovarian cancer of any stage and histological subtype, and patients with a benign ovarian mass who were referred to a highly specialized tertiary oncology unit for further assessment. Patients scheduled for pelvic surgery, involving exploratory laparotomy to determine the origin of their ovarian mass or cytoreductive surgery, were asked to collect samples prior to surgery. Patients without residual tumor/ovarian mass at time of inclusion or no possibility to collect cytological or urine samples prior to surgery were excluded from participation. Patients diagnosed with a borderline tumor were also excluded to focus on the most distinct tumor types in this feasibility stage (*i.e.,* benign and malignant ovarian masses). Patients of which not all three paired sample types (*i.e.,* cervical scrape, cervicovaginal self-sample, and urine) were available were not excluded.

Control urine samples were obtained from the URIC biobank, including healthy women without any prior cancer diagnosis within the last five years. Control cervicovaginal self-samples and cervical scrapes were collected from high-risk human papillomavirus (hrHPV)-negative women. Both were retrieved from leftover material of the Dutch national cervical cancer screening program coordinated by the Dutch National Institute for Public Health and the Environment (RIVM).

To verify the discriminatory power of the methylation assays and concordance of copy number profiles, formalin-fixed paraffin-embedded (FFPE) and fresh frozen high grade serous ovarian cancer (HGSOC) tissue samples were retrieved from the Pathology archives of Amsterdam UMC, locations AMC and VUmc, Amsterdam, The Netherlands. FFPE normal fallopian tube tissues were collected from patients undergoing a hysterectomy for the treatment of benign endometrial conditions.

### Sample collection, processing, DNA extraction, and bisulfite modification

The sample collection, processing, DNA extraction, and bisulfite modification procedures were carried out as described previously for cervical^8, 25^ and endometrial cancer.^10, 19^ A detailed description is provided in the Supplemental Methods. Briefly, urine and cervicovaginal self-samples were collected at home and clinician-taken cervical scrapes were collected before surgery. Urine was centrifuged and separated into two fractions: the urine supernatant and the urine sediment. Both fractions and the remaining full void urine were stored for further analysis. Following DNA extraction, up to 250 ng of DNA was subjected to bisulfite modification.

### DNA methylation analysis by quantitative methylation-specific PCR

Methylation levels of the *C2CD4D* (gene-ID: 100191040), *CDO1* (gene-ID: 1036), *GALR1* (gene-ID: 2587), *GHSR* (gene-ID: 2693), *MAL* (gene-ID: 4118), *NRN1* (gene-ID: 51299), *PRDM14* (gene-ID: 63978), *SST* (gene-ID: 6750), and *ZIC1* (gene-ID: 7545) genes were measured by quantitative methylation-specific polymerase chain reactions (qMSP). Methylation markers were multiplexed to assess the methylation levels of three genes (1: *GHSR/SST/ZIC1*, 2: *CDO1/MAL/PRDM14*, 3: *C2CD4D/GALR1/NRN1*) and a reference gene (*ACTB*, gene-ID: 60) within the same reaction. Methylation analysis of *CDO1*, *GALR1*, *GHSR*, *MAL*, *SST*, *PRDM14*, and *ZIC1* was performed as described previously^10, 18, 19^ with a shortened amplicon size of *ACTB*, *MAL* and *ZIC1* to facilitate methylation detection in fragmented urinary DNA. Assays targeting *C2CD4D* and *NRN1* were designed based on gene loci discovered and validated by others.^17, 21^ Primer and probe information is provided in Supplemental Table 1. Reaction conditions, instrument identifications, and thermocycling parameters are described in the Supplemental Methods. Double-stranded gBlocks™ Gene Fragments (Integrated DNA Technologies) containing the target amplicons and H_2_O were taken along in each run as positive and negative control, respectively. Sample quality and sufficient input was ensured by excluding samples with a *ACTB* quantification cycle (Cq) ≥ 32. Methylation levels were calculated relative to *ACTB* levels by the comparative Cq method: 2 ^ -(Cq marker – Cq ACTB) x 100.^26^

All qMSP assays were designed, multiplexed and optimized according to parameters described earlier.^27^ Target specificity was validated *in silico* (BLAST). Correct amplicon size was verified by agarose gel electrophoresis. Analytical validation was performed using a dilution series of bisulfite treated methylated DNA from the SiHa cell line (100, 50, 10, 5, 1, 0.5%) within the range of 20 to 0.1 ng (Supplemental Table 2). The discriminatory power of each assay was verified by comparing methylation marker levels in tissue samples of ovarian cancer patients with those measured in normal fallopian tube tissue.

### Shallow whole-genome sequencing

Urine cell-free DNA (cfDNA) extracted from urine supernatant samples of ovarian cancer patients was further characterized by shallow whole-genome sequencing (∼1x coverage). The cfDNA was quantified and analyzed using a Cell-free DNA ScreenTape assay of the Agilent 4200 TapeStation System (Agilent) for quality control before sequencing. Sequencing libraries of the first pilot series of urine supernatant DNA were prepared using the ThruPLEX Plasma-seq Kit (Takara Bio, Mountain View, CA, USA) for whole-genome sequencing according to manufacturers’ instructions. The remaining samples were prepared using the NEBNext® Enzymatic Methyl-seq (EM-seq) Kit (NEB, Ipswich, MA, USA). EM-seq was performed according to manufacturers’ guidelines for standard insert libraries with 14 PCR cycles. Libraries were quantified and quality checked using the D1000 ScreenTape Analysis Assay (Agilent) before pooling. Paired-end 150 base pair (bp) libraries were pooled in equimolar amounts and sequenced on a NovaSeq6000 (Illumina) (GenomeScan, Leiden). The processing of sequencing data and subsequent analysis of SCNA and cfDNA fragmentation patterns are provided in the Supplemental Methods. Shallow whole-genome sequencing of paired FFPE primary tumor tissue was performed to verify copy number profile concordance and is also described in the Supplemental Methods.

### Statistical analysis

Methylation levels were expressed as ^2log-^transformed Cq ratios and presented in violin plots. Tissue methylation levels were compared between two groups using the non-parametric Mann-Whitney U test. Methylation levels of each gene in the remaining sample types were compared between healthy controls and patients diagnosed with a benign or malignant ovarian mass using the Kruskal-Wallis test. In case of a significant Kruskal-Wallis test (*p*<0.05), this was followed by post-hoc testing of 1) healthy controls versus malignant ovarian masses, and 2) benign versus malignant ovarian masses using the Mann-Whitney U test with Bonferroni correction.

The correlation between methylation levels of each DNA methylation marker between paired samples of patients diagnosed with ovarian cancer was assessed using Spearman’s rank correlation. Correlation coefficient *r* was defined as very weak (*r* = 0.00–0.19), weak (*r* = 0.20–0.39), moderate (*r* = 0.40–0.59), strong (*r* = 0.60–0.79), or very strong (*r* = 0.80–1.00) and displayed in correlation matrices.

Fragment size profiles were visualized by density plots and analyzed by comparing cfDNA reads of healthy controls and ovarian cancer patients with low (<5%) and high (≥5%) tumor fractions.

Data was collected using Castor EDC and analyzed using R (version 4.0.3 with packages: cowplot, corrplot, dplyr, ggplot, ggpubr, and rstatix). *P*-values are two-sided and considered statistically significant when *p*<0.05.

## RESULTS

### Study population

A total of 428 samples of 164 participants were analyzed within this study. Samples were prospectively collected from 54 patients undergoing pelvic surgery at a tertiary oncology center because of a highly suspicious ovarian mass. Twenty-nine women were diagnosed with ovarian cancer and 25 with a benign ovarian mass. For comparison, 110 unpaired samples of healthy age-matched controls were collected. Sample types included clinician-taken cervical scrapes (control n=40, benign n=22, malignant n=24), cervicovaginal self-samples (control n=40, benign n=24, malignant n=28), full void urine (control n=30, benign n=25, malignant n=28), urine supernatant (control n=29, benign n=25, malignant n=29), and urine sediment (control n=30, benign n=25, malignant n=29). Clinical characteristics of study participants are summarized in Table 1.

**Table 1:** Clinical characteristics of study participants.

|  | n | % | Age: median (IQR) |
| --- | --- | --- | --- |
| **<u>Ovarian cancer:</u>** | 29 |  | 59 (56 - 67) |
| **Histology** |  |  |  |
| Serous carcinoma | 22 | 75.9% |  |
| Low-grade | 4 |  |  |
| High-grade | 18 |  |  |
| Clear cell carcinoma, high-grade* | 3 | 10.3% |  |
| Carcinosarcoma, high-grade | 2 | 6.9% |  |
| Endometrioid carcinoma, low-grade | 1 | 3.4% |  |
| Mucinous carcinoma, low-grade | 1 | 3.4% |  |
| **Stage (FIGO 2014)** |  |  |  |
| IIB | 5 | 17.2% |  |
| IIC | 1 | 3.4% |  |
| IIIA | 5 | 17.2% |  |
| IIIB | 4 | 13.8% |  |
| IIIC | 12 | 41.4% |  |
| IV | 2 | 6.9% |  |
| **<u>Benign ovarian mass:</u>** | 25 |  | 62 (54 - 69) |
| **Histology** |  |  |  |
| Serous cystadeno(fibro)ma | 8 | 32.0% |  |
| Mucinous cystadenoma | 6 | 24.0% |  |
| Fibroma | 4 | 16.0% |  |
| Endometriosis cyst | 4 | 16.0% |  |
| Mature teratoma | 3 | 12.0% |  |
| **<u>Healthy controls:</u>** | 110 |  |  |
| **Sample type** |  |  |  |
| Urine | 30 |  | 60 (53 - 74) |
| Cervicovaginal self-sample | 40 |  | 60 (60 - 60) |
| Clinician-taken cervical scrape | 40 |  | 60 (60 - 60) |

\*Including one mixed clear cell and low-grade endometrioid carcinoma.

### DNA methylation levels are elevated in cervical scrapes and urine samples of women with ovarian masses

The discriminatory power of qMSP assays was verified in tissue, in which all markers showed clear significant differences when comparing methylation levels in normal fallopian tube (n=22) with HGSOC (n=35) tissues (*p*<0.0001; Supplemental Figure 1, Mann-Whitney U).

The feasibility of ovarian cancer detection in urine by methylation analysis was evaluated by testing nine methylation markers in full void (*i.e.,* unfractionated) urine, urine supernatant, and urine sediment of healthy controls and patients diagnosed with a benign or malignant ovarian mass (Figure 1, Supplemental Figure 2-4). When comparing healthy controls with ovarian cancer patients, three markers showed a significant discrimination in full void urine (*C2CD4D, p*=0.008*; CDO1, p*=0.022*; MAL, p*=0.008, Mann-Whitney U), one in urine supernatant (*MAL, p=*0.001) and one in urine sediment (*GHSR, p=*0.018, Mann-Whitney U). Benign and malignant masses revealed comparably high methylation levels for most methylation markers, except for *GHSR. GHSR* showed significantly elevated methylation levels in the urine sediment of ovarian cancer patients (*p*=0.024, Mann-Whitney U; Figure 1, Supplemental Figure 4).

**Figure 1:**
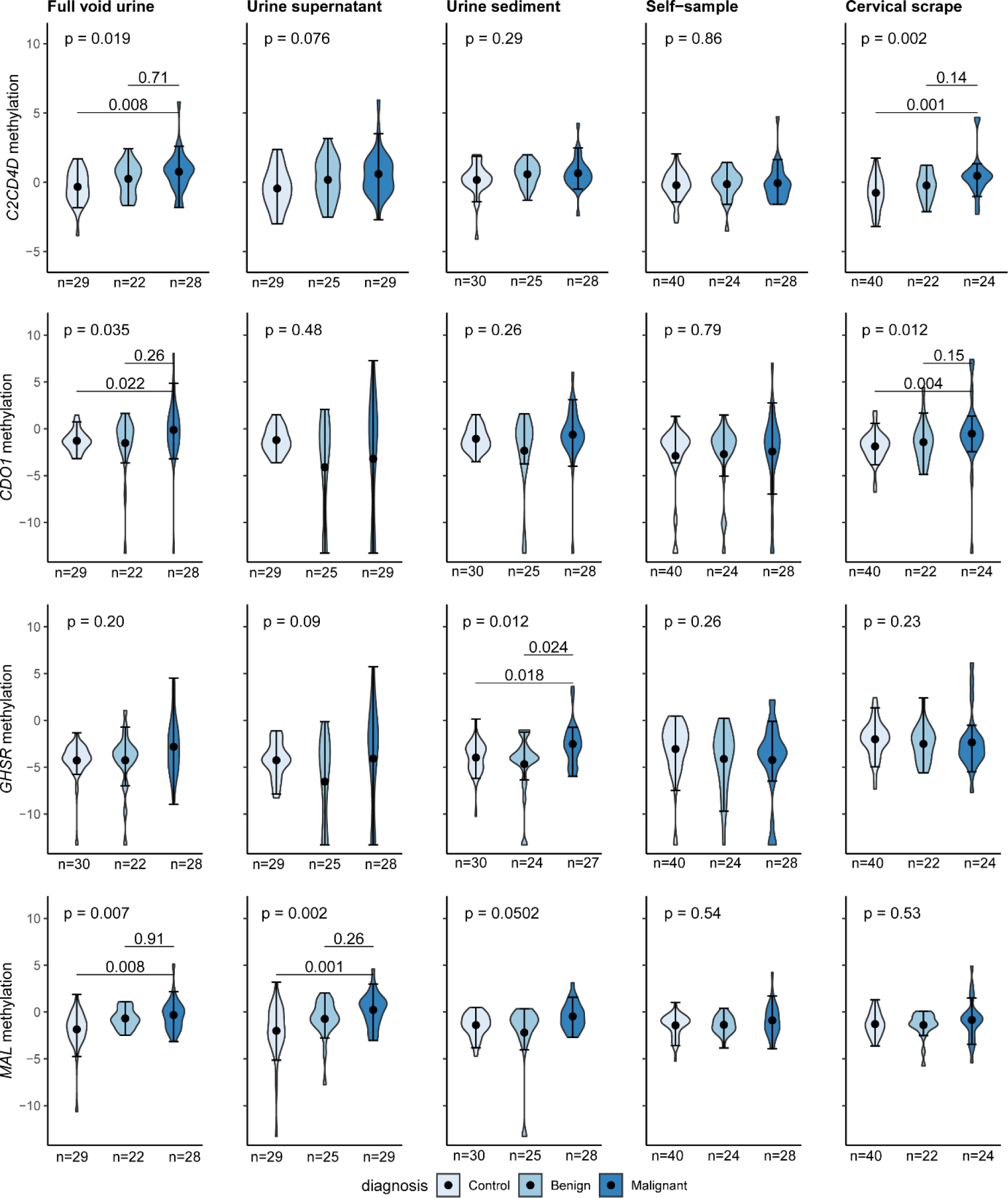
Methylation levels of most discriminating markers *C2CD4D*, *CDO1*, *GHSR*, and *MAL* in full void (unfractionated) urine, urine supernatant, urine sediment, cervicovaginal self-samples, and clinician-taken cervical scrapes of healthy controls and patients diagnosed with a benign or malignant ovarian mass. Methylation levels are expressed by 2log-transformed Cq ratios and bold circles represent medians.

Similarly, the feasibility of ovarian cancer detection in cervicovaginal self-samples and clinician-taken cervical scrapes by methylation analysis was assessed by testing the same methylation markers. While methylation levels of two markers were significantly increased in clinician-taken cervical scrapes of ovarian cancer patients as compared to controls (*C2CD4D*, *p*=0.001; *CDO1, p=*0.004, Mann-Whitney U), benign and malignant ovarian masses could not be distinguished using these markers (Figure 1, Supplemental Figure 5). None of the markers were significantly elevated in cervicovaginal self-samples when comparing these groups (Figure 1, Supplemental Figure 6).

Numbers were insufficient to compare methylation levels between different histological subtypes and stages.

### DNA methylation levels are correlated between paired cervical scrapes and urine samples

DNA methylation levels of genes significantly discriminating between healthy and malignant in cervical scrapes and urine (*i.e., C2CD4D, CDO1, GHSR, MAL*) were compared between paired samples to assess their correlation (Supplemental Figure 7). Paired cervical scrapes and urine were available for 23 ovarian cancer patients. Individual markers in full void urine correlated moderately to strongly with urine supernatant (*r* = 0.52-0.61) and urine sediment (*r* = 0.67-0.76). The full void urine showed the best correlation with cervical scrapes (*r* = 0.42-0.59), while a weak correlation was observed between the urine supernatant and cervical scrapes (*r* = 0.33-0.45).

### Copy number aberrations are detectable in urine cell-free DNA

The presence of ovarian cancer-derived DNA in the urine was verified by analyzing a subset of 25 urine supernatant samples of ovarian cancer patients (n=23) and healthy controls (n=2) by shallow whole-genome sequencing. Sequencing yielded a sufficient read count for all samples (median mapped paired read count of 55,133,492). Shallow whole-genome sequencing coverage and quality statistics per urine sample are provided in Supplemental Table 3. Aberrant genome-wide copy number profiles were found in 4 out of 23 sequenced urine supernatant samples of ovarian cancer patients (Figure 2, Supplemental Figure 8). Copy number profile concordance between urine and the primary tumor tissue was verified for these cases (Supplementary Figure 8).

**Figure 2:**
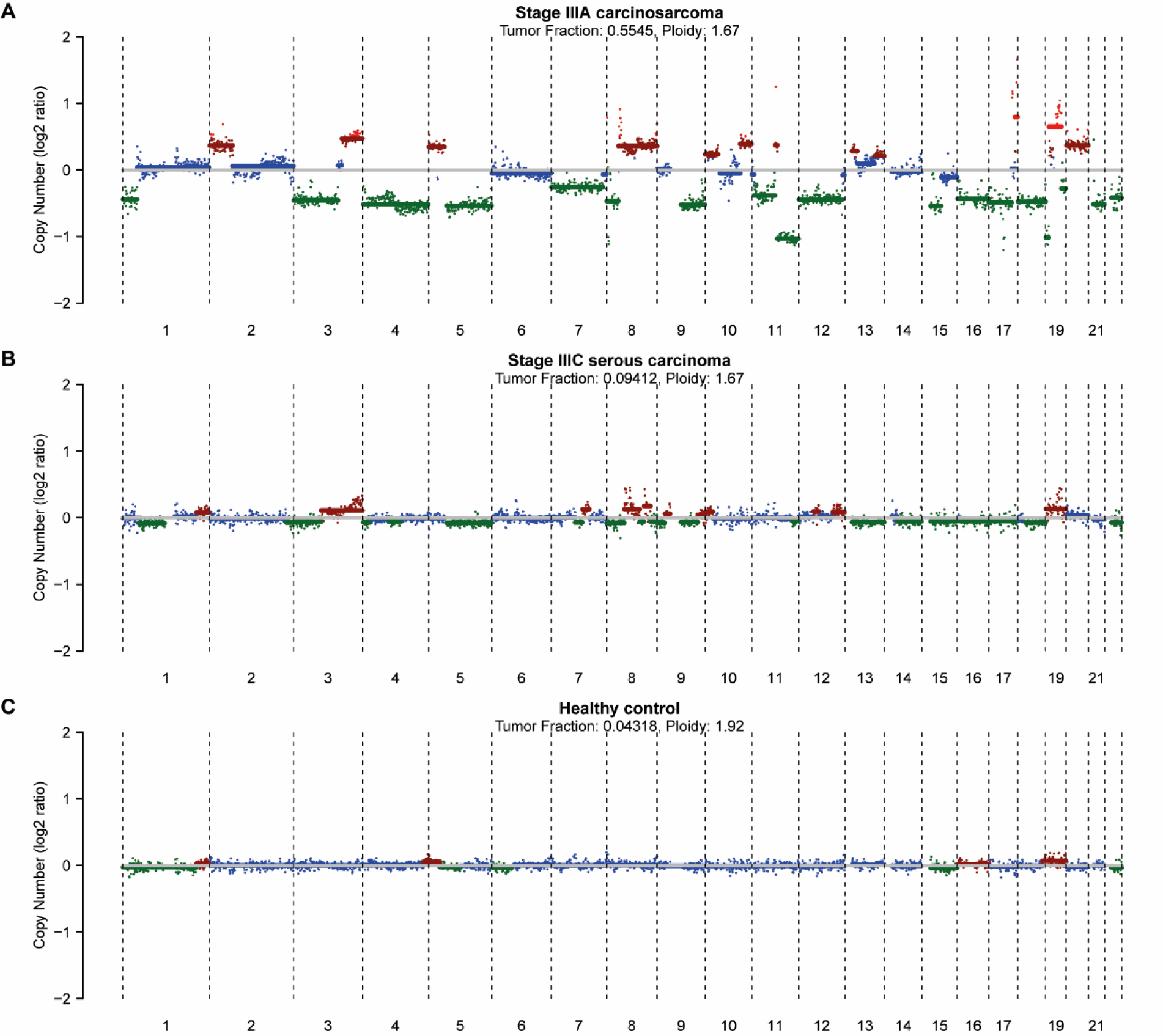
Illustrative examples of genome-wide somatic copy number profiles of urine supernatant samples collected from patients with a stage IIIA carcinosarcoma (**A**), stage IIIC serous carcinoma (**B**), and a healthy control (**C**). Estimated ploidy and tumor fraction are listed at the top of the plot. The y-axis depicts the log2 tumor to normal ratio.

The patient with the highest tumor fraction also showed the highest methylation levels of *MAL* in the urine supernatant (Supplemental Figure 9). Additionally, fragment size distributions were analyzed by comparing cfDNA reads of healthy controls and ovarian cancer patients with low and high tumor fractions. Cancer samples with a high tumor fraction (n=4) revealed a shorter modal fragment size of 80 bp as compared to 111 bp in cancer samples with a low tumor fraction (n=19) and controls (n=2; Supplemental Figure 10).

## DISCUSSION

Both elevated methylation levels of a subset of markers and SCNA were detected in home-collected urine samples of ovarian cancer patients by targeted qMSP assays and shallow whole-genome sequencing, respectively. Urine is truly non-invasive and unlocks at home collection of liquid biopsy to reduce in-person visits. Yet, an important finding was that methylation levels in benign cases were similarly high, presenting a challenge for the development of clinically useful tests.

While we tested for methylation markers described and also by us verified to be associated with ovarian cancer, it was found that when tested in our patient-friendly sample types most of these did not distinguish benign from malignant ovarian masses. Only *GHSR* demonstrated slightly increased methylation levels in the urine sediment. Benign ovarian masses included in this study were highly suspicious for malignancy according to current triage methods (>40% risk of malignancy using the IOTA adnex model) as samples were collected in a tertiary oncology unit. Half of the included patients in our cohort were ultimately diagnosed with a benign ovarian mass, underlining that current triage for referral to tertiary oncology care is suboptimal. The majority of previous studies only included benign controls for methylation marker discovery in tissue but not during marker validation in plasma, as recently reviewed by Terp *et al.* ^15^, or benign controls were not age-matched to cancers.^21^ Similarly, studies on ovarian cancer detection in cervical scrapes did not include benign controls.^16, 17^ The inclusion of age-matched patients diagnosed with benign and malignant ovarian masses is essential to accurately assess the clinical value of DNA methylation testing for ovarian cancer detection.

The presence of ovarian cancer-derived DNA in the urine is currently underexplored. So far, only Valle *et al*. reported on the detection of somatic mutation profiles and *HIST1H2BB/MAGI2* promoter methylation in a small paired series of ascites, blood, tissue, urine, and vaginal swabs of HGSOC patients.^28^ Their data on two patients revealed that methylation levels in urinary cfDNA correlated stronger with tissue than with blood, indicating the potential of urine-based ovarian cancer detection. Unfortunately, the diagnostic potential of ovarian cancer detection in urine could not be determined in the study of Valle *et al*. as no control samples were included.

In our study, different urine fractions were systematically compared to explore whether a preferred urine sample type for ovarian cancer detection exists. Full void urine most likely contains both genomic and cfDNA, whereas the urine sediment is enriched for genomic DNA and the urine supernatant for transrenally excreted cfDNA.^29^ This assumption is confirmed by the strong correlation for *CDO1* between cervical scrapes and urine sediment, while cervical scrapes and urine supernatant correlated weakly to moderately. Most methylation markers significantly differentiated between healthy controls and ovarian cancer patients in the full void urine (3/12), followed by urine supernatant (1/12), and the urine sediment (1/12). These outcomes suggest that tumor-derived methylation signals can originate from genomic DNA as well as transrenally excreted cfDNA. Yet, larger samples sizes are needed to determine whether a preferred urine sample type for methylation analysis exists.

In the present study, genes with elevated methylation levels in HGSOC tissue, were not always measurable in urine. Our qMSP assays were designed to facilitate the detection of methylation in small DNA fragments present in the urine as shown in our previous studies.^8, 10, 12^ Yet, the current assays may not reach the limit of detection needed for the low tumor-derived methylation signals. Nucleic acids that are released from the bladder epithelium may further dilute the ovarian cancer signal in urine. Another explanation for the absence of tumor-derived methylation signals of some genes in the urine could be linked to the origin of urinary cfDNA. Urine cfDNA is described to be even shorter as compared to plasma cfDNA (modal size of 82 vs. 167 basepairs) ^30^. Differences in fragmentation patterns between plasma and urine are likely caused by Dnase1 cleavage activity in the urine and high concentrations of urea and salt that affect histone-DNA binding ^31^. Histone-bound DNA is more protected against degradation as compared to DNA that is not histone-bound ^32^. Hypothetically, hypermethylated regions of interest that are not histone-bound could be further degraded and become unmeasurable. We partly accounted for this by including methylation markers with proven diagnostic value in plasma in our selection (*i.e., C2CD4D*^21, 22^, *CDO1*^22^), which both appeared suitable for ovarian cancer detection in urine.

Clear SCNA profiles harboring common chromosomal gains (*e.g.,* 1q, 3q, 7q, 8q) and losses (*e.g.,* 17p, 19q, 22q) could be obtained from four urine supernatant samples of ovarian cancer patients, verifying the presence of tumor-derived DNA in the urine.^33^ Furthermore, a focal amplification at chromosome 19 was identified in the urine of one patient with stage IIIA serous carcinoma, which is a clinically relevant alteration that has previously been described in a subgroup of serous ovarian cancers.^34^ Aneuploidy was detected previously in cervical scrape samples of ovarian cancer patients using the PapSEEK test ^16^. We also observed shorter fragment sizes in urine supernatant samples with a high tumor fraction, which is another indication for the presence of tumor-derived DNA in the urine, as shown previously in urine samples of glioma patients ^30^.

Given the feasibility of ovarian cancer detection in cervical scrapes by DNA methylation analysis^14, 17^, similar findings were expected for self-collected cervicovaginal samples. While *C2CD4D* and *CDO1* distinguished healthy versus malignant in cervical scrapes, none of the markers showed elevated methylation levels in cervicovaginal self-samples. Our findings are in line with those of van Bommel *et al*. who reported that mutation analysis in cervicovaginal self-samples of ovarian cancer patients was not feasible.^35^ None of the pathogenic mutations found in surgical specimens could be detected in cervicovaginal self-samples. Ovarian cancer signals might be more diluted in cytological specimens collected from areas further away from the ovaries. This was also observed for the PapSEEK test, which detected 45% of ovarian cancers when using intrauterine sampling (Tao brush) as compared to 17% when using endocervical sampling (Pap brush).^16^

Nevertheless, considering our relatively small sample size, we do not exclude the use of cervicovaginal self-samples for ovarian cancer detection yet. The optimization of pre-analytical factors, such as increased input of original sample or improved DNA isolation methods, could enhance the ovarian cancer signal in vaginal samples. Alternatively, a non-tumor DNA driven approach could be useful for ovarian cancer detection in cervicovaginal self-samples, as recently described by Barrett *et al*.^36^ Their signature consisted of epigenetic differences in cervical cells and allowed ovarian cancer detection in cervical scrapes with an area under the receiver operating characteristic curve value of 0.76. Larger cohort studies, such as the Screenwide study ^37^, will provide further insight into the use of cervicovaginal self-samples for ovarian cancer detection.

Strengths of this study include the collection of a unique paired sample series of both patients diagnosed with a benign ovarian mass and with a malignant ovarian tumor, covering most histological subtypes. Moreover, urine and cervicovaginal self-samples were collected from home to assess the feasibility and potential of home-based sampling for ovarian cancer. The successful sequencing of urine cfDNA of ovarian cancer patients provides opportunities for future (epi)genome profiling using short- or long-read sequencing technologies. Although we have demonstrated the potential diagnostic value of urine for ovarian cancer, this study is limited by still relatively low sample numbers and the lack of early-stage cancers (≤ FIGO stage 2A). Given the heterogeneous nature of benign and malignant ovarian masses, larger sample series are needed to conclude on the clinical applicability of home-collected cervicovaginal self-samples and urine for ovarian cancer detection. Furthermore, direct comparisons with paired plasma samples using DNA-based and other molecular biomarkers (*e.g.,* HE4) would be informative for future studies.

This study supports limited existing data on ovarian cancer detection in cervical scrapes by DNA methylation analysis. Moreover, it provides first proof of concept that urine yields increased methylation levels of ovarian cancer-associated genes and contains ovarian cancer-derived DNA as demonstrated by SCNA analysis. Our findings support continued research into urine biomarkers for ovarian cancer detection and highlight the importance of including benign ovarian masses in future studies. Molecular biomarker testing in patient-friendly samples could facilitate earlier ovarian cancer detection and triage women presenting with an ovarian mass to manage specialist referral. Yet, further studies investigating alternative urine (methylation) biomarkers are warranted to develop a clinically useful test.

## Glossary

cfDNA: cell-free DNA
Cq: quantification cycle
FFPE: formalin-fixed paraffin-embedded
HGSOC: high grade serous ovarian cancer
hrHPV: high-risk human papillomavirus
qMSP: quantitative methylation-specific PCR
SCNA: somatic copy number aberrations

## Acknowledgements

The authors thank Dr. Poell, Dr. Kasius, and Dr. Mom for the useful discussions and support during funding acquisition. The authors also thank Dr. Y. Kim, J.M.P Egthuijsen, N. Hogervorst, and N. Evander for technical assistance and A. Koch for help during sample collection.

## Author contributions

**BW:** Funding acquisition, resources, data curation, formal analysis, investigation, visualization, methodology, writing-original draft. **MS**: Resources, data curation, writing-review and editing. **MB:** Conceptualization, supervision, funding acquisition, validation, writing-review and editing, project administration. **YB**: Investigation, data curation, methodology, writing-review and editing. **RH:** Conceptualization, resources, writing-review and editing. **CL:** Resources, writing-review and editing. **FD**: Resources, writing-review and editing. **YP**: Investigation, writing-review and editing. **FM**: Resources, formal analysis, visualization, methodology, writing-review and editing. **NM:** Resources, formal analysis, visualization, methodology, writing-review and editing. **NT:** Conceptualization, supervision, funding acquisition, resources, validation, writing-review and editing, project administration. **RS:** Conceptualization, supervision, funding acquisition, validation, writing-original draft, project administration.

## Ethics statement

All patients participating in the SOLUTION1 study were 18 years or older and signed informed consent before sample collection. Ethical approval was obtained by the Medical Ethical Committee of the VU University Medical Center for the use of samples collected within the SOLUTION1 study (METc: 2016.213, Trial registration ID: NL56664.029.16), samples stored in the URIC biobank (TcB 2018.657), and samples archived in the biobank containing leftover material of the Dutch national cervical cancer screening program (TcB 2020.245). The Code of Conduct for Responsible Use of Left-over Material of the Dutch Federation of Biomedical Scientific Societies was adhered for the use of tissue specimen.

## Data and code availability

The sequencing dataset generated and analyzed during the current study is available in the European Genome-Phenome Archive repository, under accession number EGAD00001010848. The DNA methylation dataset generated and analyzed during this study is available from the corresponding author on reasonable request.

