## Supplemental Methods for "Molecular analysis for ovarian cancer detection in patient-friendly samples"

### Sample collection and processing

Urine and cervicovaginal self-samples were collected at home for which all participants received a package including materials needed for collection and transport. Participants were instructed to collect urine before the cervicovaginal self-sample. Cervicovaginal self-samples were collected according to the provided user manual using the Evalyn® brush (Rovers Medical Devices, Oss, The Netherlands), which is a clinically validated self-sampling method.[1] Urine was collected in 3x30 mL tubes containing the storage buffer Ethylenediaminetetraacetic acid (EDTA; final concentration 40mM) to preserve nucleic acids during transport. Clinician-taken cervical scrapes were collected prior to surgery using a Cervex-Brush (Rovers Medical Devices) and directly placed in 10 mL Thinprep PreservCyt medium (Hologic, Marlborough, MA, US). Samples were sent to the Pathology department of Amsterdam UMC, location VUmc, within 72 hours by regular mail and processed directly after arrival.

Urine was processed as described in our previously validated processing and storage protocol.[2] Briefly, a total of 15 mL of urine was centrifuged at 3000g for 10 min to separate the urine into two fractions: the urine supernatant and urine sediment. Both fractions and the remaining full void (*i.e.* unfractionated) urine were stored at -20°C. Cytological samples were processed as described previously for cervical[3] and endometrial cancer.[4] Cervicovaginal self-samples were stored in 1.5 mL ThinPrep PreservCyt medium upon arrival. Cervicovaginal self-samples and cervical scrapes were stored at 4°C.

Formalin-fixed paraffin-embedded (FFPE) and fresh frozen tissue specimens were consecutively sectioned of which the first and last sections were Hematoxylin and Eosin (H&E) stained for histopathological review by a pathologist to confirm the presence of ovarian cancer or normal fallopian tube tissue.

### DNA extraction and bisulfite modification

DNA from full void urine (30 mL patients diagnosed with ovarian mass; 40 mL controls), urine sediment (15 mL original volume), and urine supernatant (15 mL) was extracted as described previously.[5, 6] In short, both full void urine and urine supernatant were isolated with the Quick DNA urine kit (Zymo Research, Irvine, CA, US) and urine sediment using the DNA mini and blood mini kit (Qiagen, Hilden,

Germany). DNA from cervicovaginal self-samples and clinician-taken cervical scrapes was isolated as described before[3], using the NucleoMag 96 Tissue kit (Machery-Nagel) and a Microlab Star robotic system (Hamilton, Germany). DNA of FFPE tissue samples was isolated using the QIAamp DNA FFPE tissue kit (Qiagen, Hilden, Germany). DNA of fresh frozen tissue samples was isolated using the DNeasy Blood & Tissue kit (Qiagen). DNA yield was quantified using a NanoDrop 1000 (Thermo Fisher Scientific, Waltham, MA, US). Up to 250 ng of extracted DNA was subjected to bisulfite modification using the EZ DNA Methylation Kit (Zymo Research) to convert unmethylated cytosines. All procedures were performed according to manufacturers' guidelines.

**Reaction conditions and instrument identifications of quantitative methylation-specific PCR**

Up to 50 ng of modified DNA was mixed with Epitect Multiplex PCR Mastermix (Qiagen, Venlo, Netherlands), 2.5-5.0 µM of each primer, and 5.0-10.0 µM of each hydrolysis probe in a total volume of 12.5 µl. Thermocycling conditions were: 95°C for 5 minutes, 45 cycles at 95°C for 15 seconds, 59/60/63°C for 1 minute, and 72°C for 1 minute. Quantitative methylation-specific PCR (qMSP) assays were performed using a ViiA7 real-time PCR-system (Applied Biosystems, Foster City, CA, USA) or an ABI-7500 real-time PCR-system (Applied Biosystems, Waltham, MA, US) for *GHSR/SST/ZIC1*. The qMSP data was analyzed with manual thresholds and automatic baseline settings using QuantStudio™ Real-Time PCR Software (v. 1.6.1) and 7500 Software (v. 2.3).

**Analysis of somatic copy number aberrations and cell-free DNA fragmentation patterns**

Processing of the sequencing data was performed by a pipeline controlled by Snakemake (v. 7.14.0). In brief, sequencing adapters and indexes were trimmed by the bbduk.sh (v. 38.79) [https://sourceforge.net/projects/bbmap/] in paired mode with parameters 'ktrim=r k=23 mink=11 hdist=1' and the adapter reference dataset provided with the software. Trimmed non-converted samples were mapped to the GRCh38 human genome assembly (GeneBank accession: GCA_000001405.28) using bwa mem (v. 0.7.17) [https://github.com/lh3/bwa]. Enzymatically converted reads were mapped to the same assembly using biscuit (v. 1.0.2.20220113) [https://huishenlab.github.io/biscuit/]. For both non-converted and converted samples, reads with a mapping quality lower than 5, unmapped reads, secondary mappings, chimeric and PCR duplicates were filtered using samtools (v. 1.12) [https://github.com/samtools/samtools] and sambamba markdup (v. 0.8.1) [https://lomereiter.github.io/sambamba/]. Reads passing the filtering step were submitted for somatic copy number aberrations (SCNA) analysis and tumor fraction estimation using the ichorCNA software (v. 0.3.2.0)[7] using default settings, except the use of an in-house panel-of-normals from shallow whole-genome sequencing, setting the non-tumor fraction parameter restart values to c(0.95,0.99,0.995,0.999). The tumor fraction with the highest log likelihood was reported. Fragmentation patterns of urine cfDNA for both non-converted and converted samples were analyzed by retrieving the fragment sizes of the trimmed and filtered reads using picard CollectInsertSizeMetrics (v. 2.22.2) with HISTOGRAM_WIDTH=1000 [https://gatk.broadinstitute.org/hc/en-us].

Shallow whole-genome sequencing for the analysis of SCNA in paired FFPE primary tumor tissue was performed as described previously with a few adaptations.[8] A PCR-free VeriSeq workflow (Illumina) was used in combination with the Illumina NextSeq 500. Sequence reads were aligned to the GRCh38 human genome assembly using bwa mem (v. 0.7.17). PCR duplicates (marked by Picard v. 2.20.8), as well as low-quality reads (MAPQ < 37), were filtered out using samtools (v. 0.1.1830). Reads passing the filtering step were submitted for SCNA analysis using ichorCNA software as described for urine samples.
