## Supplemental Figure 1 - 10 for "Molecular analysis for ovarian cancer detection in patient-friendly samples"

### SUPPLEMENTAL FIGURES

#### Table of contents


| | |
|---|---|
| **Supplemental Figure 1** | DNA methylation levels of *C2CD4D, CDO1, GALR1, GHSR, MAL, NRN1, PRDM14, SST,* and *ZIC1* in high grade serous ovarian cancer (n=35) and normal fallopian tube tissue (n=22). DNA methylation levels are shown by 2log-transformed Cq ratios. Violin plots represent medians with lower and upper quartile and range whiskers. A *p*-value of 0.05 was considered statistically significant. ****: $p < 0.0001$. Cq = quantification cycle; HGSOC = high grade serous ovarian cancer. |
| **Supplemental Figure 2** | DNA methylation levels of *C2CD4D, CDO1, GALR1, GHSR, MAL, NRN1, PRDM14, SST,* and *ZIC1* in full void (*i.e.* unfractionated) urine of healthy controls (n=30), and women diagnosed with a benign (n=27) or high stage malignant ovarian mass (n=28). DNA methylation levels are shown by 2log-transformed Cq ratios. Violin plots represent medians with lower and upper quartile and range whiskers. A *p*-value of <0.05 was considered statistically significant. Cq = quantification cycle. |
| **Supplemental Figure 3** | DNA methylation levels of *C2CD4D, CDO1, GALR1, GHSR, MAL, NRN1, PRDM14, SST,* and *ZIC1* in urine supernatant of healthy controls (n=29), and women diagnosed with a benign (n=27) or high stage malignant ovarian mass (n=29). DNA methylation levels are shown by 2log-transformed Cq ratios. Violin plots represent medians with lower and upper quartile and range whiskers. A *p*-value of <0.05 was considered statistically significant. Cq = quantification cycle. |
| **Supplemental Figure 4** | DNA methylation levels of *C2CD4D, CDO1, GALR1, GHSR, MAL, NRN1, PRDM14, SST,* and *ZIC1* in urine sediment of healthy controls (n=30), and women diagnosed with a benign (n=27) or high stage malignant ovarian mass (n=29). DNA methylation levels are shown by 2log-transformed Cq ratios. Violin plots represent medians with lower and upper quartile and range whiskers. A *p*-value of <0.05 was considered statistically significant. Cq = quantification cycle. |
| **Supplemental Figure 5** | DNA methylation levels of *C2CD4D, CDO1, GALR1, GHSR, MAL, NRN1, PRDM14, SST,* and *ZIC1* in clinician-collected cervical scrapes of healthy controls (n=40), and women diagnosed with a benign (n=23) or high stage malignant ovarian mass (n=24). DNA methylation levels are shown by 2log-transformed Cq ratios. Violin plots represent medians with lower and upper quartile and range whiskers. A *p*-value of <0.05 was considered statistically significant. Cq = quantification cycle. |
| **Supplemental Figure 6** | DNA methylation levels of *C2CD4D, CDO1, GALR1, GHSR, MAL, NRN1, PRDM14, SST,* and *ZIC1* in self-collected cervicovaginal samples of healthy controls (n=40), and women diagnosed with a benign (n=25) or high stage malignant ovarian mass (n=28). Violin plots represent medians with lower and upper quartile and range whiskers. DNA methylation levels are shown by 2log-transformed Cq ratios. A *p*-value of <0.05 was considered statistically significant. Cq = quantification cycle. |
| **Supplemental Figure 7** | The Spearman correlation coefficients (*r*) of methylation markers *C2CD4D, CDO1, GHSR,* and *MAL* between paired samples of 23 women diagnosed with ovarian cancer. The Spearman correlation coefficient was calculated based on 2log-transformed Cq ratios. Circle color and size indicate the degree of correlation (*i.e*., the larger and darker the circle, the more correlation). |
| **Supplemental Figure 8** | Genome-wide SCNA profiles of matched urine and FFPE primary tumor tissue The log2 tumor to normal ratio is depicted on the y-axis and the chromosomal position on the x-axis. Computed using ichorCNA software. SCNA = somatic copy number aberrations. FFPE = formalin-fixed paraffin-embedded, SCNA = somatic copy number aberrations. |
| **Supplemental Figure 9** | Scatter plot indicating the relation between *MAL* methylation levels and the tumor fraction as estimated by ichorCNA in urine supernatant samples. *MAL* methylation levels are shown by 2log-transformed Cq ratios. *MAL* was the most discriminating marker between urine supernatant samples of healthy controls and ovarian cancer patient and therefore plotted against the tumor fraction. The patient with the highest tumor fraction in urinary cfDNA also showed the highest *MAL* methylation, as seen in the upper right part of the plot. Cq = quantification cycle. |
| **Supplemental Figure 10** | Fragment size distributions for cfDNA reads of urine supernatant samples from healthy controls (n=2) and ovarian cancer patients with a low (<5%, n=19) and high (≥5%, n=4) tumor fraction determined from shallow whole-genome sequencing. The cfDNA with a high tumor fraction revealed a shorter modal fragment size (80 bp) than cfDNA with a low tumor fraction and controls (111 bp). |

**Supplemental Figure 1:** DNA methylation levels of *C2CD4D, CDO1, GALR1, GHSR, MAL, NRN1, PRDM14, SST,* and *ZIC1* in high grade serous ovarian cancer (n=35) and normal fallopian tube tissue (n=22). DNA methylation levels are shown by 2log-transformed Cq ratios. Violin plots represent medians with lower and upper quartile and range whiskers. A *p*-value of 0.05 was considered statistically significant. ****: $p < 0.0001$. Cq = quantification cycle; HGSOC = high grade serous ovarian cancer.

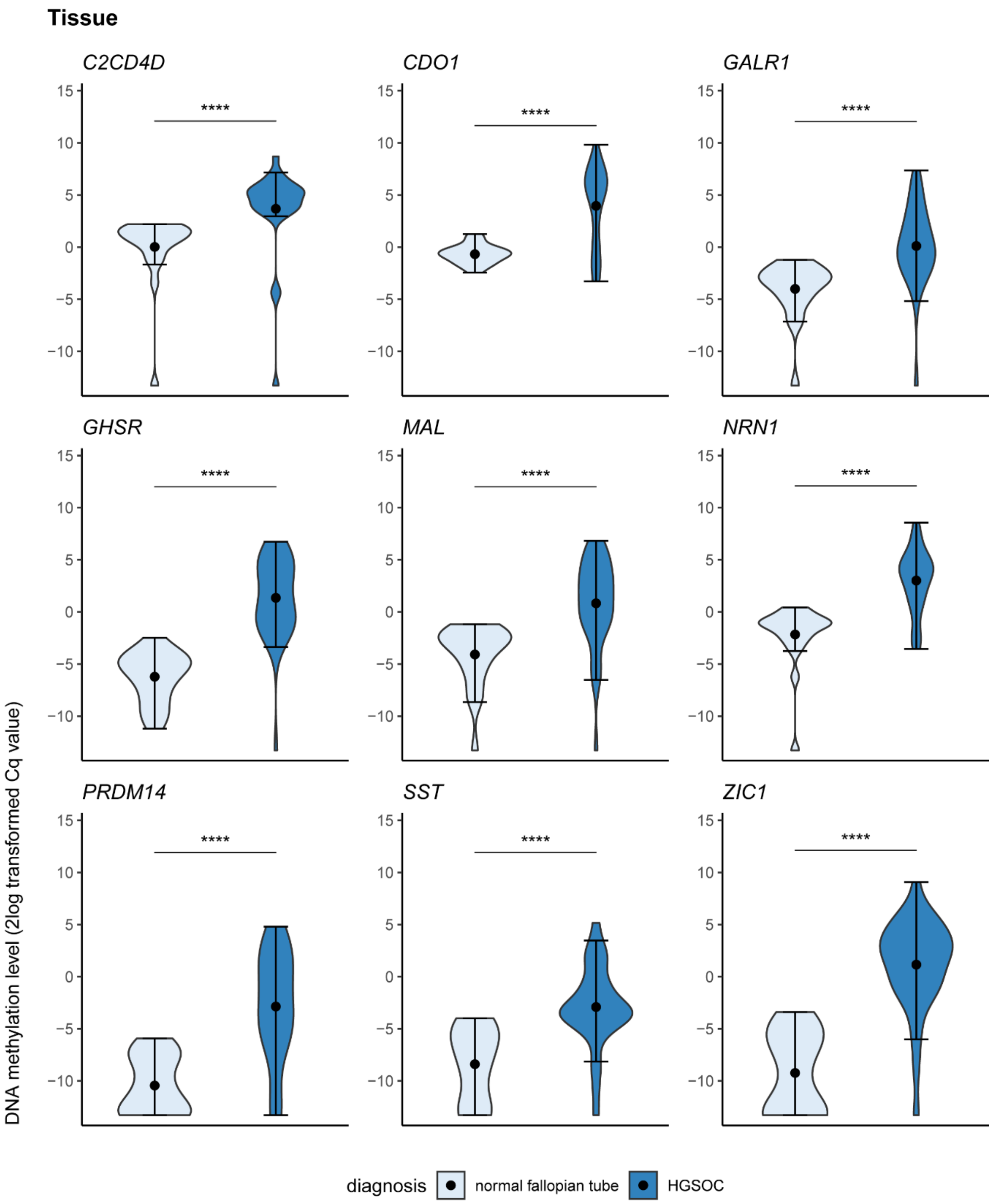

**Supplemental Figure 2:** DNA methylation levels of *C2CD4D, CDO1, GALR1, GHSR, MAL, NRN1, PRDM14, SST*, and *ZIC1* in full void (*i.e.* unfractionated) urine of healthy controls (n=30), and women diagnosed with a benign (n=27) or high stage malignant ovarian mass (n=28). DNA methylation levels are shown by 2log-transformed Cq ratios. Violin plots represent medians with lower and upper quartile and range whiskers. A *p*-value of <0.05 was considered statistically significant. Cq = quantification cycle.

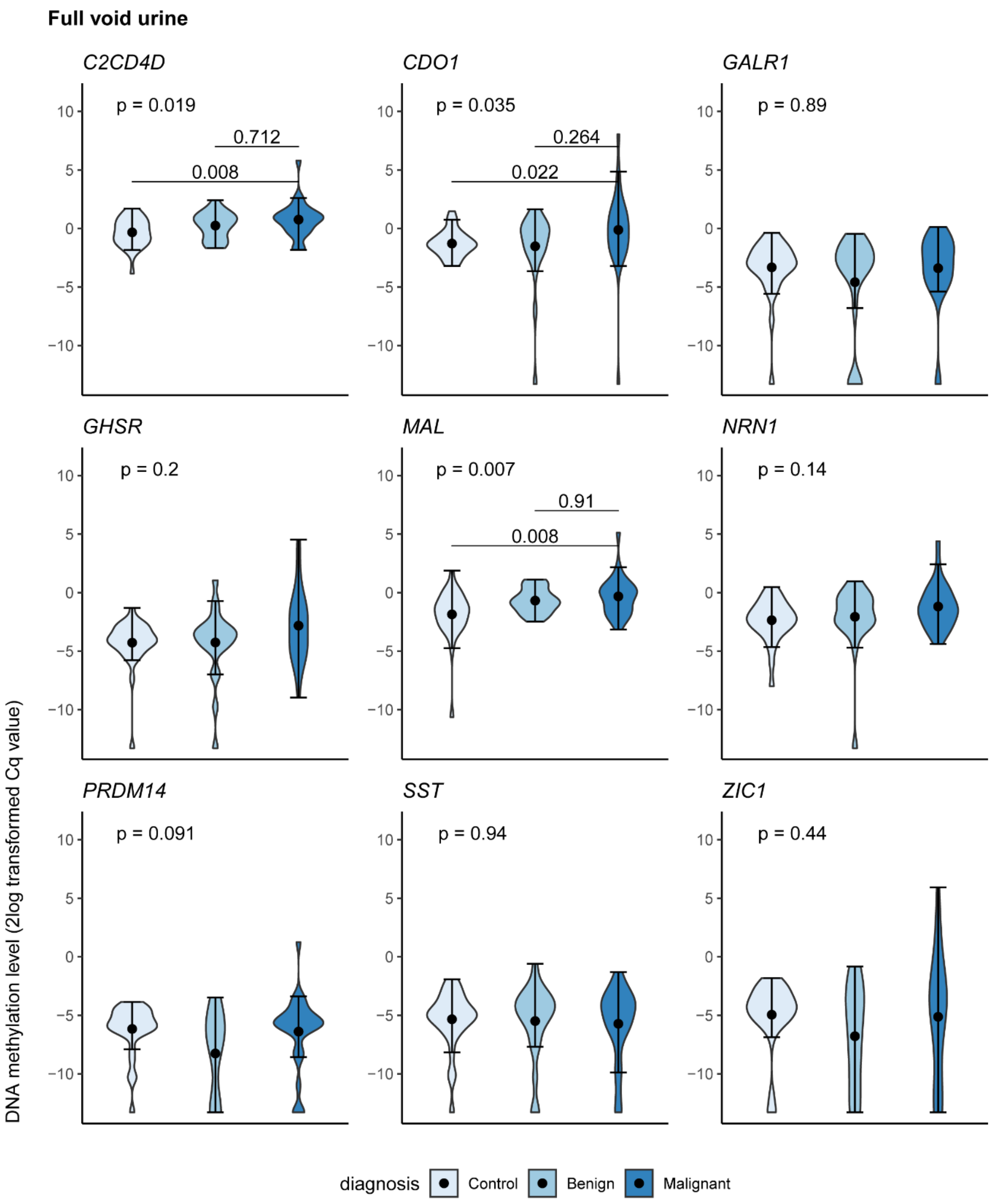

**Supplemental Figure 3:** DNA methylation levels of *C2CD4D, CDO1, GALR1, GHSR, MAL, NRN1, PRDM14, SST,* and *ZIC1* in urine supernatant of healthy controls (n=29), and women diagnosed with a benign (n=27) or high stage malignant ovarian mass (n=29). DNA methylation levels are shown by 2log-transformed Cq ratios. Violin plots represent medians with lower and upper quartile and range whiskers. A *p*-value of <0.05 was considered statistically significant. Cq = quantification cycle.

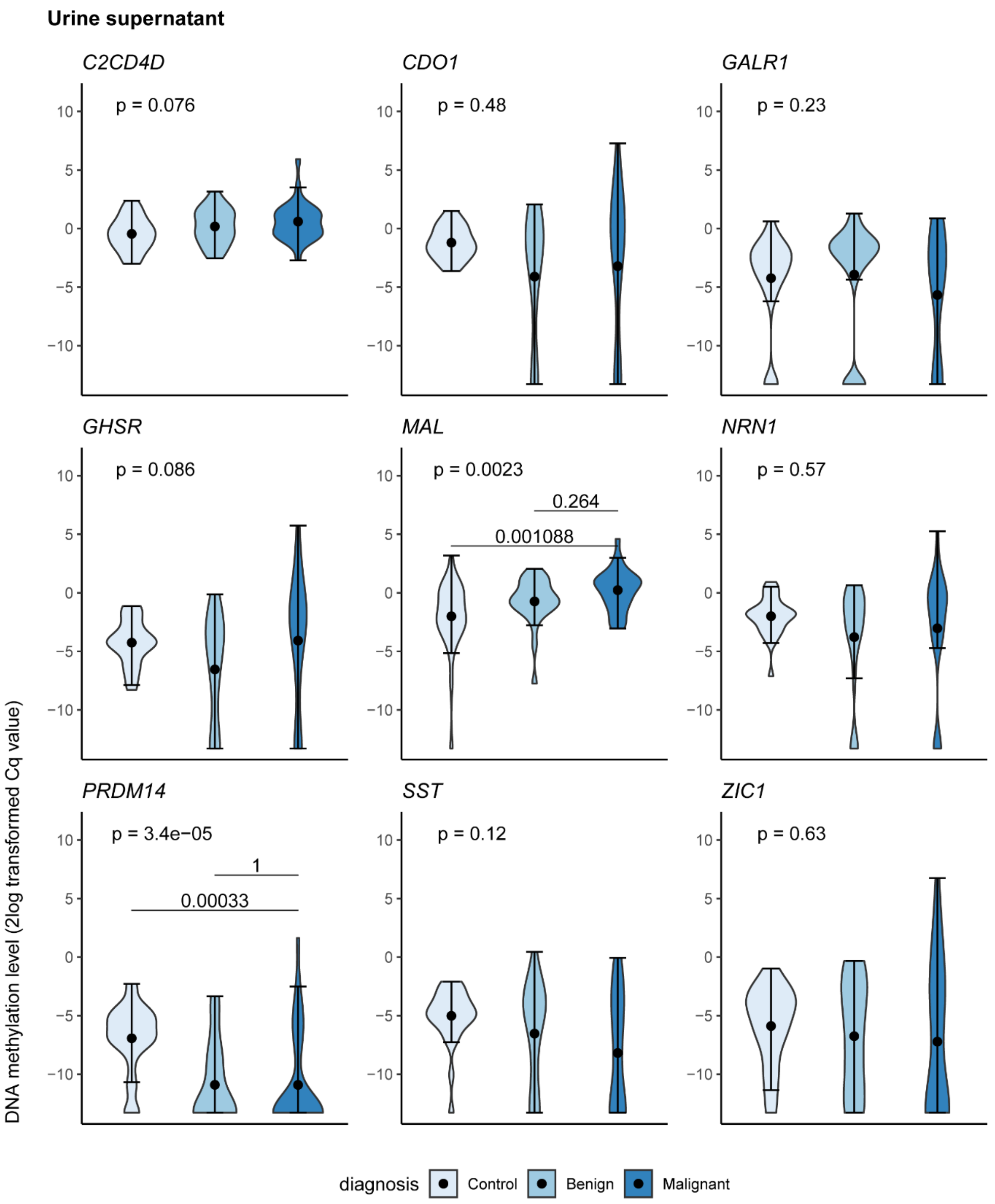

**Supplemental Figure 4:** DNA methylation levels of *C2CD4D, CDO1, GALR1, GHSR, MAL, NRN1, PRDM14, SST,* and *ZIC1* in urine sediment of healthy controls (n=30), and women diagnosed with a benign (n=27) or high stage malignant ovarian mass (n=29). DNA methylation levels are shown by 2log-transformed Cq ratios. Violin plots represent medians with lower and upper quartile and range whiskers. A *p*-value of <0.05 was considered statistically significant. Cq = quantification cycle.

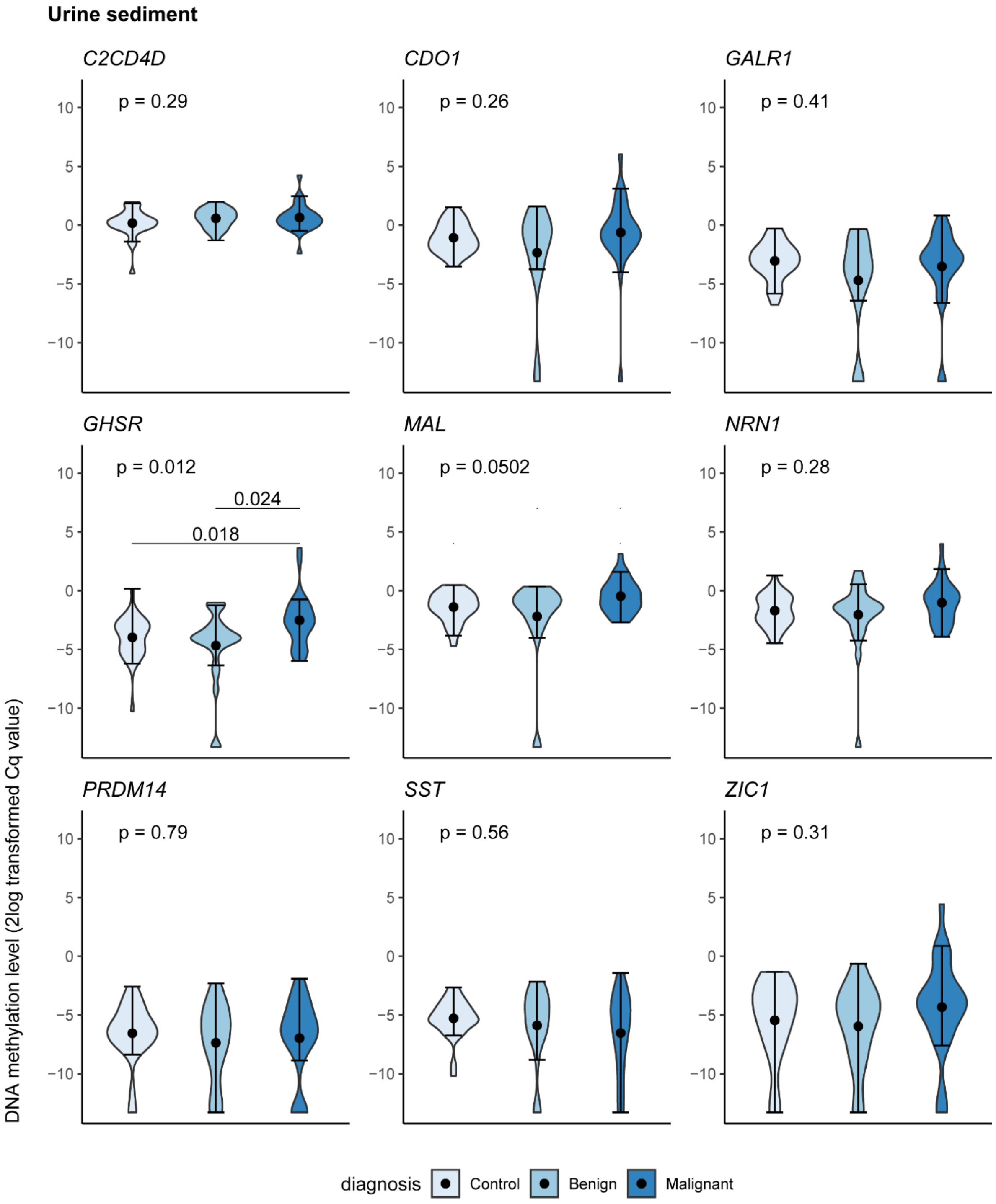

**Supplemental Figure 5:** DNA methylation levels of *C2CD4D, CDO1, GALR1, GHSR, MAL, NRN1, PRDM14, SST,* and *ZIC1* in clinician-taken cervical scrapes of healthy controls (n=40), and women diagnosed with a benign (n=23) or high stage malignant ovarian mass (n=24). DNA methylation levels are shown by 2log-transformed Cq ratios. Violin plots represent medians with lower and upper quartile and range whiskers. A *p*-value of <0.05 was considered statistically significant. Cq = quantification cycle.

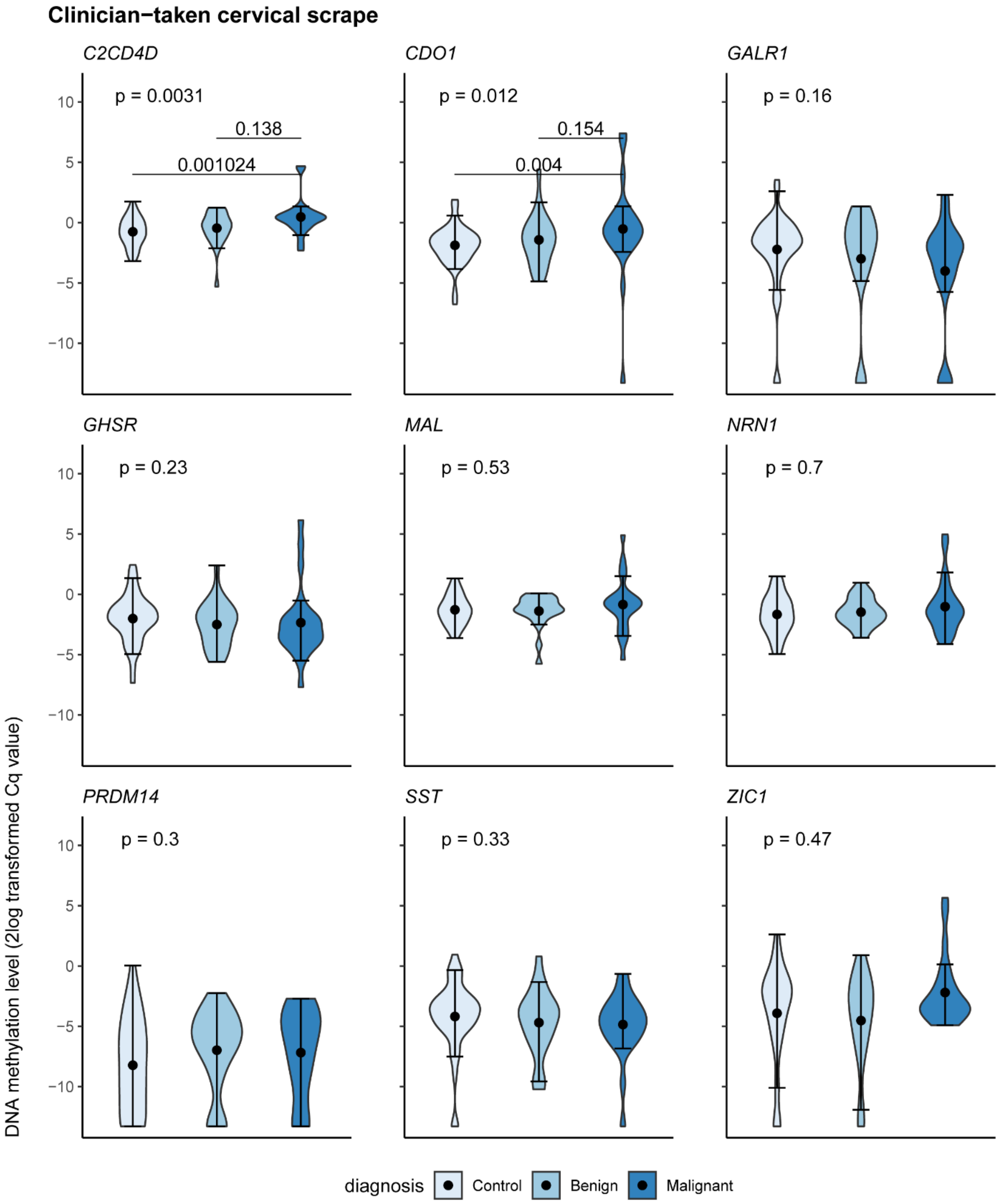

**Supplemental Figure 6**: DNA methylation levels of *C2CD4D, CDO1, GALR1, GHSR, MAL, NRN1, PRDM14, SST,* and *ZIC1* in self-collected cervicovaginal samples of healthy controls (n=40), and women diagnosed with a benign (n=25) or high stage malignant ovarian mass (n=28). Violin plots represent medians with lower and upper quartile and range whiskers. DNA methylation levels are shown by 2log-transformed Cq ratios. A *p*-value of <0.05 was considered statistically significant. Cq = quantification cycle.

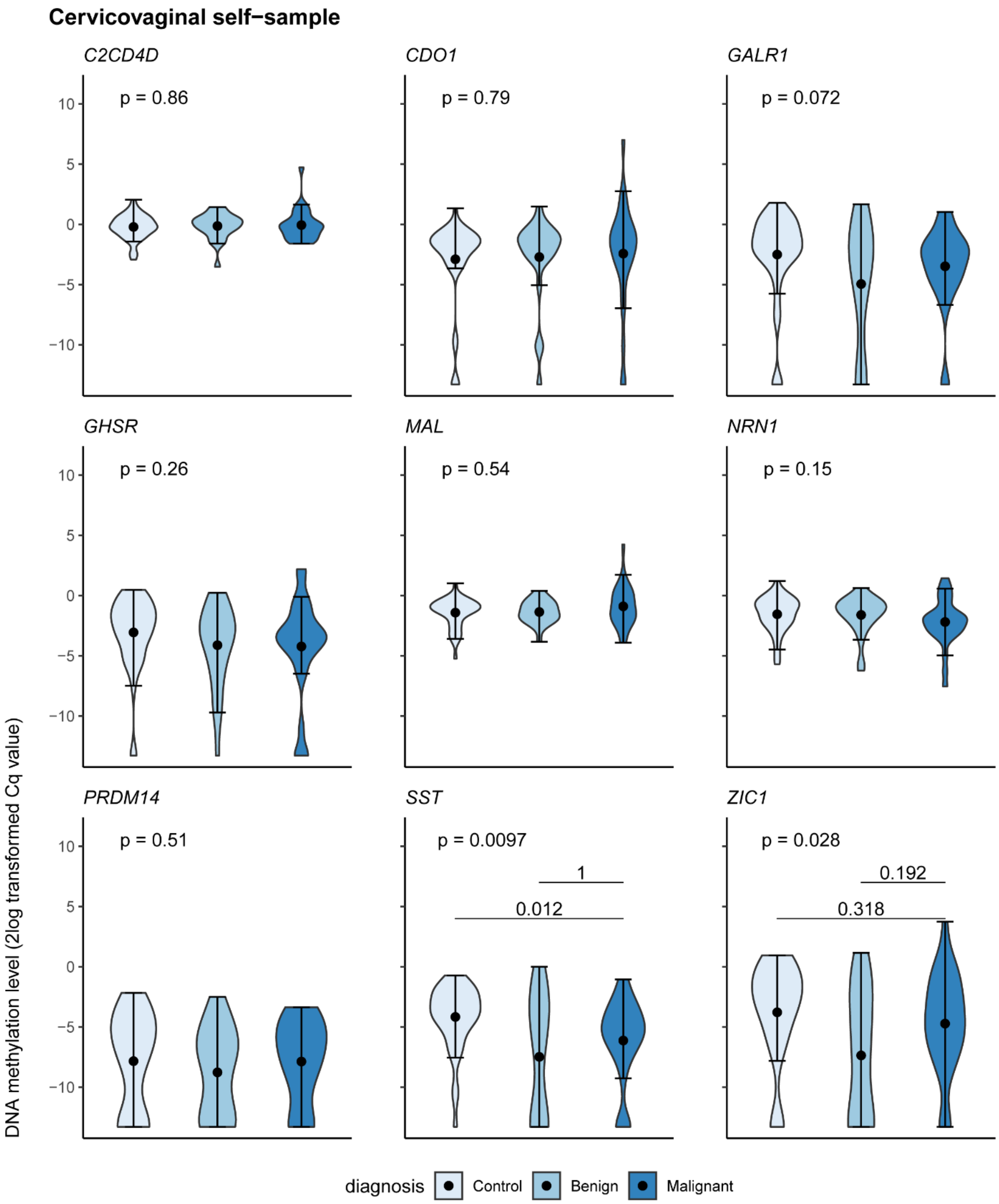

**Supplemental Figure 7**: The Spearman correlation coefficients (*r*) of methylation markers *C2CD4D, CDO1, GHSR,* and *MAL* between paired samples of 23 women diagnosed with ovarian cancer. The Spearman correlation coefficient was calculated based on 2log-transformed Cq ratios. Circle color and size indicate the degree of correlation (*i.e.*, the larger and darker the circle, the more correlation).

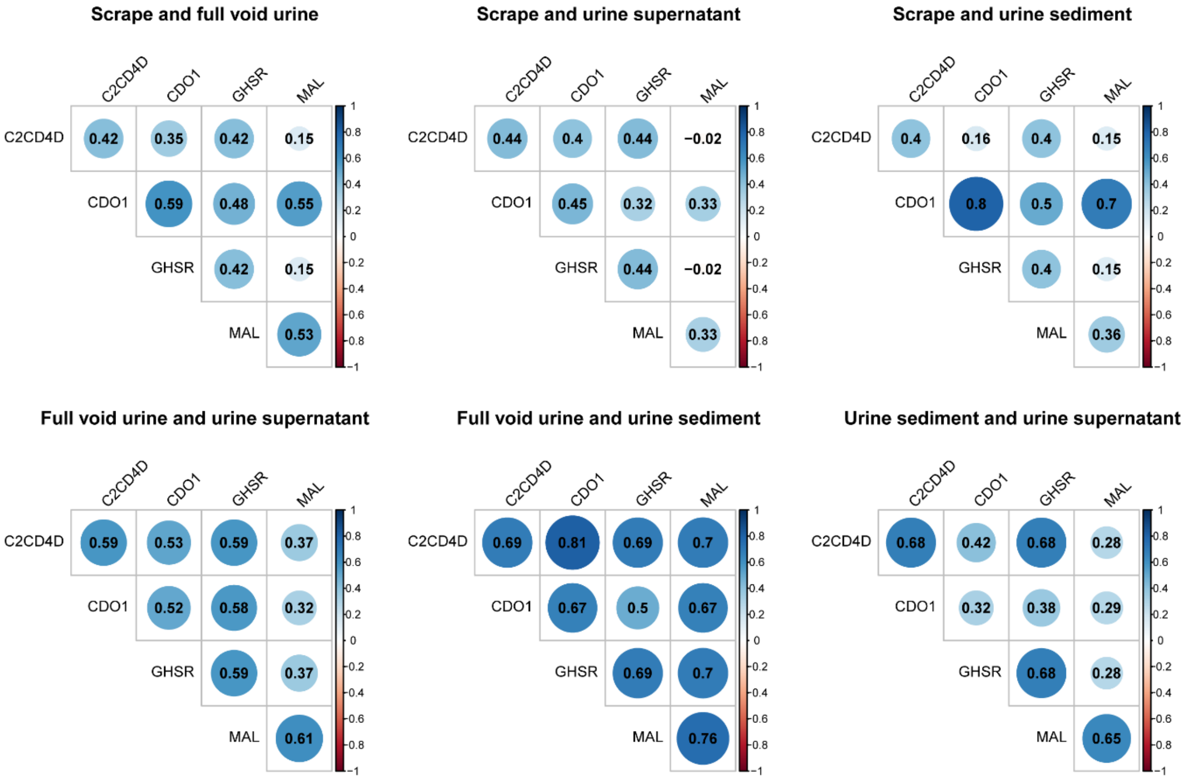

**Supplemental Figure 8:** Genome-wide SCNA profiles of matched urine and FFPE primary tumor tissue

The log2 tumor to normal ratio is depicted on the y-axis and the chromosomal position on the x-axis. Computed using ichorCNA software. SCNA = somatic copy number aberrations. FFPE = formalin-fixed paraffin-embedded, SCNA = somatic copy number aberrations.

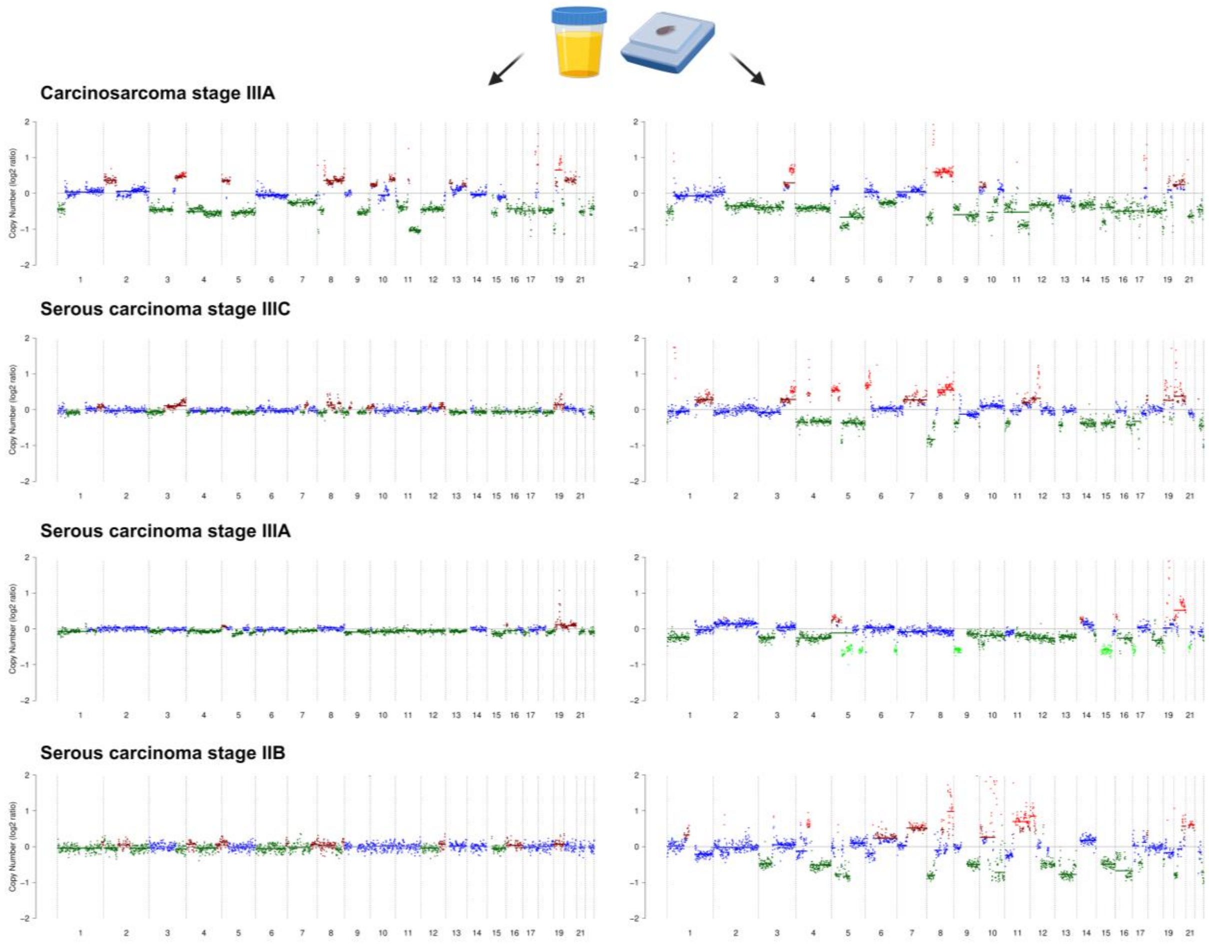


Created with BioRender.com.

**Supplemental Figure 9:** Scatter plot indicating the relation between *MAL* methylation levels and the tumor fraction as estimated by ichorCNA in urine supernatant samples. *MAL* methylation levels are shown by 2log-transformed Cq ratios. *MAL* was the most discriminating marker between urine supernatant samples of healthy controls and ovarian cancer patient and therefore plotted against the tumor fraction. The patient with the highest tumor fraction in urinary cfDNA also showed the highest *MAL* methylation, as seen in the upper right part of the plot. Cq = quantification cycle.

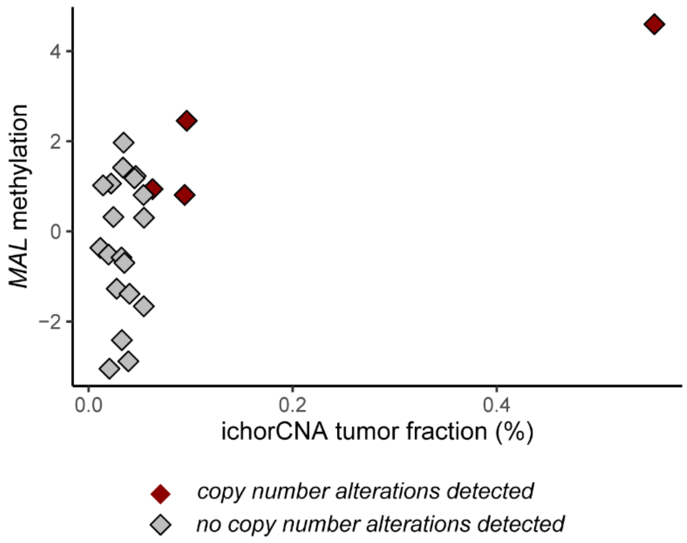

**Supplemental Figure 10:** Fragment size distributions for cfDNA reads of urine supernatant samples from healthy controls (n=2) and ovarian cancer patients with a low (<5%, n=19) and high (≥5%, n=4) tumor fraction determined from shallow whole-genome sequencing. The cfDNA with a high tumor fraction revealed a shorter modal fragment size (80 bp) than cfDNA with a low tumor fraction and controls (111 bp).

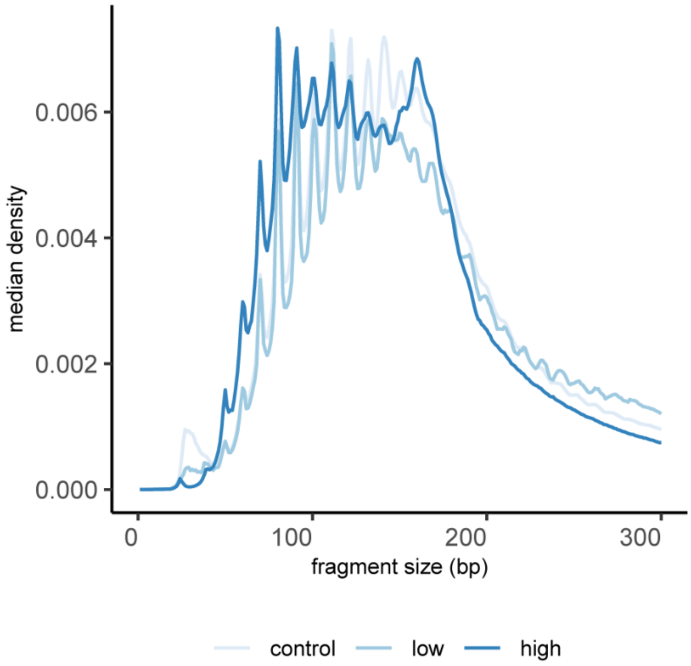
